# Synthesising artificial patient-level data for Open Science - an evaluation of five methods

**DOI:** 10.1101/2020.10.09.20210138

**Authors:** Michael Allen, Andrew Salmon

**Affiliations:** University of Exeter Medical School & NIHR South West Peninsula Applied Research Collaboration (ARC)

## Abstract

**Background:** Open science is a movement seeking to make scientific research accessible to all, including publication of code and data. Publishing patient-level data may, however, compromise the confidentiality of that data if there is any significant risk that data may later be associated with individuals. Use of synthetic data offers the potential to be able to release data that may be used to evaluate methods or perform preliminary research without risk to patient confidentiality.

**Methods:** We have tested five synthetic data methods:

1. A technique based on *Principal Component Analysis (PCA)* which samples data from distributions derived from the transformed data.
2. *Synthetic Minority Oversampling Technique, SMOTE* which is based on interpolation between near neighbours.
3. *Generative Adversarial Network, GAN*, an artificial neural network approach with competing networks - a *discriminator* network trained to distinguish between synthetic and real data., and a *generator* network trained to produce data that can fool the discriminator network.
4. *CT-GAN*, a refinement of GANs specifically for the production of structured tabular synthetic data.
5. *Variational Auto Encoders, VAE*, a method of encoding data in a reduced number of dimensions, and sampling from distributions based on the encoded dimensions.

Two data sets are used to evaluate the methods:

1. The *Wisconsin Breast Cancer data set*, a histology data set where all features are continuous variables.
2. A *stroke thrombolysis pathway data set*, a data set describing characteristics for patients where a decision is made whether to treat with clot-busting medication. Features are mostly categorical, binary, or integers.

Methods are evaluated in three ways:

1. The ability of synthetic data to train a logistic regression classification model.
2. A comparison of means and standard deviations between original and synthetic data.
3. A comparison of covariance between features in the original and synthetic data.

**Results:** Using the Wisconsin Breast Cancer data set, the original data gave 98% accuracy in a logistic regression classification model. Synthetic data sets gave between 93% and 99% accuracy. Performance (best to worst) was SMOTE > PCA > GAN > CT-GAN = VAE. All methods produced a high accuracy in reproducing original data means and stabdard deviations (all R-square > 0.96 for all methods and data classes). CT-GAN and VAE suffered a significant loss of covariance between features in the synthetic data sets.

Using the Stroke Pathway data set, the original data gave 82% accuracy in a logistic regression classification model. Synthetic data sets gave between 66% and 82% accuracy. Performance (best to worst) was SMOTE > PCA > CT-GAN > GAN > VAE. CT-GAN and VAE suffered loss of covariance between features in the synthetic data sets, though less pronounced than with the Wisconsin Breast Cancer data set.

**Conclusions:** The pilot work described here shows, as proof of concept, that synthetic data may be produced, which is of sufficient quality to publish with open methodology, to allow people to better understand and test methodology. The quality of the synthetic data also gives promise of data sets that may be used for screening of ideas, or for research project (perhaps especially in an education setting).

More work is required to further refine and test methods across a broader range of patient-level data sets.

## 1 Introduction

Open science is a movement seeking to make scientific research accessible to all ^1^. This includes not only open publication of scientific papers, but provision of code (using open source software), and underlying data. Publishing patient-level data may, however, compromise the confidentiality of that data if there is any significant risk that data may later be associated with individuals.

If we are to publish analysis or models, such as machine learning models, with data we therefore need a means of producing data that contains all the features of the original data but does not present any significant risk to patient confidentiality. We may take two approaches to solving this problem. Firstly we may add noise to the data to sufficiently protect anonymity; this approach is used in the *differential privacy* method ^2^. A second approach is to try to produce synthetic data that is a reasonable facsimile of the original data, but that does not directly recreate original data points.

In this paper we present experiments with five different methods of producing synthetic data:

1. A method based on *Principal Component Analysis, PCA*, a classical statistical method for dimensionality reduction ^3^. Data is transformed into *k* orthogonal dimensions. We use this approach to create synthetic data by sampling from distributions for each Principal Component dimension, and transforming these sampled data points back into the original data dimension space.
2. *Synthetic Minority Oversampling Technique, SMOTE* ^4^. This is a method normally used to enhance data with extra points created by interpolation between near neighbours. Here we follow the same methodology used for data augmentation, but remove the original data points, leaving only the synthetic data points.
3. *Generative Adversarial Network, GAN* ^5^. This method relies on two adversarial artificial neural networks. A *discriminator* network is trained to distinguish between synthetic and real data. A *generator* network is trained to produce data that can fool the discriminator network. The performance of each improves as the two networks are trained in contest with each other.
4. *Conditional Tabular GAN, CT-GAN* ^6^. CT-GAN is a development of a general GAN with the aim of providing synthetic tabular data. A *conditional GAN* framework is used to help prevent *modal collapse*, a problem where a GAN may generate realistic synthetic data, but that the the population variance of the synthetic data is significantly reduced compared to the original data set.
5. *Variational Auto Encoders, VAE* ^7^. An *autoencoder* is a type of artificial neural network that encodes data in a reduced dimension space ^8^. The network is trained so that data is forced down through a layer (the *latent space* layer with fewer dimensions than the original data. Decoding layers then expand back to the original number of dimensions, and the network is trained to minimise the loss between the decoded data and the original data. Variational Auto Encoders are an adaptation to allow this framework to be used for synthetic data production, using a specialised way of regulating the network to avoid over-fitting to the original data. The latent layer is framed as a distribution for each dimension, with the loss function for training the model incorporating a penalty for low variance distributions. Synthetic data is produced by sampling values for the latent layers using the distribution parameters obtained in training of the network.

Clinical data can take various forms. Here we investigate techniques using two different data sets.

1. The *Wisconsin Breast Cancer data set* ^9^. This is a set of histology data. There are two classes: samples that come from malignant tumours and samples that come from benign tumours. All features are continuous variables. The data set contains 569 tissue samples each with 30 features.
2. A *stroke thrombolysis pathway data set* ^10^. This is a data set of clinical characteristics for patients in an acute Stroke Pathway. There are two classes: patients that receive thrombolysis (treatment to dissolve a clot in the brain), and patients that do not receive thrombolysis. The majority of features are categorical, binary, or integer values. The data set contains 1862 patients each with 50 features.

This work describes initial pilot work to test the five synthetic data methods applied to the two data sets. Methods are not fully optimised, and the two data sets while intending to represent two different types of data, do not represent all types of clinical data.

## 2 Methods

All data and code for the experiments described here may be found at https://github.com/MichaelAllen1966/synthetic_data_pilot/releases/tag/1.0.0 (DOI:10.5281/zenodo.4075288). All methods are coded in Python ^11^.

### 2.1 Synthetic methods

Methods coded are:

1. *Principal Component Analysis, PCA* ^3^, implemented using the *decomposition* package in SciKit Learn ^12^.
2. *Synthetic Minority Oversampling Technique, SMOTE* ^4^, implemented using the IMBLearn package ^13^.
3. *Generative Adversarial Network, GAN* ^5^, implemented in PyTorch ^14^
4. *Conditional Tabular GAN, CT-GAN* ^6^, implemented using the CT-GAN package (https://github.com/sdv-dev/CTGAN).
5. *Variational Auto Encoders, VAE* ^7^ implemented in TensforFlow ^15^ and Keras (https://github.com/fchollet/keras.)

Additional numerical calculations are performed using NumPy ^16^ and Pandas ^17^. Results are plotted using MatPlotLib ^18^.

All synthetic methods have been constructed to to initially produce continuous variable outputs. Non continuous outputs are generated as follows:

1. *Binary* : values of 0.5 or greater are set to 1, otherwise 0.
2. *Integer* : values are set as the rounded integer of the continuous variable. No clipping is applied.
3. *Categorical* : values are converted to one-hot encoding in the raw data. In the synthetic data the one-hot feature with the greatest value is set to 1, and all others are set to 0.

### 2.2 evaluation

For each method the synthetic method is run separately for the negative and positive class examples.

Methods are evaluated in three ways:

1. A logistic regression model (SciKit Learn ^12^) is trained using synthetic data. This is then tested against 25% of the original data (the remaining 75% of the original data is used to train another logistic regression model for comparison).
2. Means and standard deviations are compared between original and synthetic data. Coefficient of correlation of the comparison is described in the results section. Detailed plots are provided in the appendix.
3. Covariance between features is evaluated in the original and synthetic data. The pair-wise coefficient of correlation for each feature pair is compared between original and synthetic data and an overall coefficient of correlation (of the pair-wise coefficients of correlation between original and synthetic data) provided in the results. Detailed plots are provided in the appendix.

## 3 Results

### 3.1 Training a logistic regression classification model

Table 1 shows the performance of a logistic regression classification model trained on original or synthetic data (when original data is used to train the model, the model is tested on 25% of the data not used to train the model). Results are shown for accuracy (proportion of all cases identified correctly), sensitivity (proportion of positive cases identified correctly) and specificity (proportion of negative cases identified correctly). The experiment was repeated five times.

**Table 1:** Performance of original and synthetic data sets when used to train a logistic regression model, which is tested against original data. Results show mean ± sem (n=5).

| Data set | Synthesis | Accuracy | Sensitivity | Specificity |
| --- | --- | --- | --- | --- |
| Wisconsin | Original | 0.975 (0.007) | 0.944 (0.011) | 0.995 (0.004) |
|  | PCA | 0.976 (0.003) | 0.971 (0.098) | 0.980 (0.005) |
|  | SMOTE | 0.990 (0.003) | 0.985 (0.013) | 0.993 (0.002) |
|  | GAN | 0.957 (0.008) | 0.934 (0.016) | 0.968 (0.007) |
|  | CT-GAN | 0.933 (0.015) | 0.825 (0.030) | 0.998 (0.002) |
|  | VAE | 0.933 (0.009) | 0.965 (0.019) | 0.998 (0.002) |
| Stroke | Original | 0.815 (0.005) | 0.806 (0.008) | 0.821 (0.005) |
|  | PCA | 0.801 (0.010) | 0.875 (0.005) | 0.752 (0.015) |
|  | SMOTE | 0.824 (0.004) | 0.853 (0.012) | 0.804 (0.010) |
|  | GAN | 0.678 (0.012) | 0.725 (0.056) | 0.644 (0.048) |
|  | CT-GAN | 0.684 (0.034) | 0.877 (0.029) | 0.556 (0.075) |
|  | VAE | 0.664 (0.004) | 0.692 (0.031) | 0.648 (0.002) |

### 3.2 Comparison of means and standard deviations

Table 2 shows a summary of correlations between original and synthetic means and standard deviations (see appendix for detailed charts). Further detailed results are available in the Jupyter Notebooks in the on-line GitHub repository.

**Table 2:** R-squared for correlations between original and synthetic means and standard deviations (SD). Results show mean ± sem (n=5).

| Data set | Synthesis | Means |  | Standard Deviations |  |
| --- | --- | --- | --- | --- | --- |
|  |  | Negative | Positive | Negative | Positive |
| Wisconsin | PCA | 1.000 (0.000) | 1.000 (0.000) | 1.000 (0.000) | 1.000 (0.000) |
|  | SMOTE | 1.000 (0.000) | 1.000 (0.000) | 1.000 (0.000) | 1.000 (0.000) |
|  | GAN | 0.999 (0.000) | 0.999 (0.000) | 1.000 (0.000) | 0.999 (0.000) |
|  | CT-GAN | 0.985 (0.006) | 0.967 (0.010) | 0.996 (0.001) | 0.975 (0.003) |
|  | VAE | 0.998 (0.001) | 0.983 (0.007) | 0.980 (0.011) | 0.974 (0.008) |
| Stroke | PCA | 1.000 (0.000) | 1.000 (0.000) | 0.999 (0.000) | 1.000 (0.000) |
|  | SMOTE | 1.000 (0.000) | 1.000 (0.000) | 0.999 (0.000) | 0.998 (0.000) |
|  | GAN | 0.999 (0.000) | 1.000 (0.000) | 0.967 (0.004) | 0.987 (0.003) |
|  | CT-GAN | 0.998 (0.001) | 0.996 (0.001) | 0.996 (0.001) | 0.988 (0.003) |
|  | VAE | 0.998 (0.001) | 0.999 (0.000) | 0.987 (0.005) | 0.989 (0.004) |

Table 3 shows a summary of correlation coefficients between original and synthetic pair-wise feature correlation coefficients (see appendix for detailed charts).

**Table 3:** R-squared for correlation between original and synthetic pair-wise feature correlation coefficients. Results show mean ± sem (n=5).

| Data set | Synthesis | Negative | Positive |
| --- | --- | --- | --- |
| Wisconsin | PCA | 0.996 (0.000) | 0.995 (0.000) |
|  | SMOTE | 0.984 (0.002) | 0.981 (0.002) |
|  | GAN | 0.918 (0.009) | 0.832 (0.039) |
|  | CT-GAN | 0.134 (0.002) | 0.146 (0.006) |
|  | VAE | 0.535 (0.044) | 0.166 (0.038) |
| Stroke | PCA | 0.939 (0.002) | 0.934 (0.001) |
|  | SMOTE | 0.923 (0.001) | 0.910 (0.002) |
|  | GAN | 0.833 (0.006) | 0.828 (0.008) |
|  | CT-GAN | 0.388 (0.003) | 0.448 (0.003) |
|  | VAE | 0.844 (0.008) | 0.827 (0.006) |

## 4 Discussion

In the data sets we have examined here synthetic approaches based on PCA and SMOTE have the best performance overall: classification model performance is maintained, means and standard deviations of the synthetic data closely match the original data set, and covariance between features is well maintained. A standard GAN performed reasonably well in all categories. CT-GAN, however performed more poorly in the classification model training for the stroke model and, while means and standard deviations closely matched the original data set, there was a significant loss in covariance between the features. The VAE performance was similar to the standard GAN, but also suffered from some loss in covariance between features, especially in the Wisconsin Breast Cancer data set.

PCA and SMOTE currently appear the best choices for synthesising tabular patient data, but testing on more data sets is required. PCA may struggle as feature sets become larger and computation of the principal components becomes more computationally challenging.

GANs are a rapidly developing type of network. They are able to synthesise complex data including non-structured data such as images (see https://thispersondoesnotexist.com as an example of a GAN creating realistic images of people. Here we have used just a simple GAN and CT-GAN. There is potential in testing developments of the GAN approach, such as the Wasserstein GAN ^19^ which improves stability of GANs and helps prevent modal collapse where the synthetic data is realistic but from the population of the synthetic data is more limited in variance than that of the original data.

Instability in GANs is a well known phenomenon, hence the development of techniques such as Wasserstein GAN to improve stability. One practical approach may also be to train an ensemble of GANs, and choose the one that produces the highest quality synthetic data.

The relatively poor performance of the CT-GAN, especially the profound loss of the expected covariance between features in the synthetic data, was a surprise, as this method is targeted at replicating tabular data. From our results, this method should be used with caution where preservation of feature covariance is important.

The overall performance of the VAEs was similar to the GAN, except that there was some loss in covariance between features. Unlike PCA there is no requirement of the encoded reduced dimension layer to have encoded features that are orthogonal to each other, so it is perhaps not surprisingly that the VAE does not necessarily maintain feature covariance.

Whether performance of a synthetic method is sufficiently good, and which method is best, depends on the purpose of the synthetic data. Is the synthetic data to be used as an illustrative data set, or will detailed analysis be performed on it? For Open Science, the former will probably most common - the synthetic data must resemble the original data with close enough approximation that the methods and results being presented may be understood using the synthetic data. A next step up the synthetic data quality ladder is to use of synthetic data that may be made publicly available and that can be used to test ideas before an application is made for robust analysis of the original data. The final rung of the synthetic data quality ladder is when synthetic data may totally replace original data for research, with no need even to confirm results using the original data. The quality of patient-level synthetic data, from our pilot experiments, appears to be within this spectrum - easily good enough to be used to help people understand and test published methodology, and likely good enough to be used to screen ideas (e.g. in an educational research setting).

### 4.1 Limitations

Two key limitations of the work described here are:

1. We have so far used only two patient-level data sets. Those these data sets were chosen to represent different types of data, further evaluation is needed using alternative patient-level data sets.
2. We have used methods in their basic configuration. Further optimisation, or use of refined approaches, may improve on performance observed here.
3. In all our methods we trained the synthetic data engines on data from each data class separately. We have yet to evaluate the performance of machine learning classification trained on synthetic data where the class is treated as just one of the features in the data set (a single synthetic data engine would be trained and used, rather than class-based engines).

### 4.2 Further work

Further work will focus on the following areas:

1. Optimising methods and using refinements to methods (especially more advanced GAN techniques).
2. Testing on a broader range of patient-level data sets.
3. Testing the ability to produce synthetic data suitable for machine learning classification with-out the need to separately train methods on different classes of data.
4. Testing ensembles of artificial neural nets (GANs, VAEs), picking the best performing engine and testing against a separate held-back data set (for machine learning classification).

## 5 Conclusions

The pilot work described here shows, as proof of concept, that synthetic data may be produced, which is of sufficient quality to publish with open methodology, to allow people to better understand and test methodology. The quality of the synthetic data also gives promise of data sets that may be used for screening of ideas, or for research project (perhaps especially in an education setting).

## Data Availability

The code and data used in this study is available at https://github.com/MichaelAllen1966/synthetic_data_pilot/releases/tag/1.0.0

## Appendices

### A Wisoconsin Breast Cancer data set

Figures 1 to 10 show a) a comparison of means and standard deviations between the original and synthetic data sets, and b) correlation between all features in original and synthetic data sets. The figures show five synthetic runs for each method.

**Figure 1:**
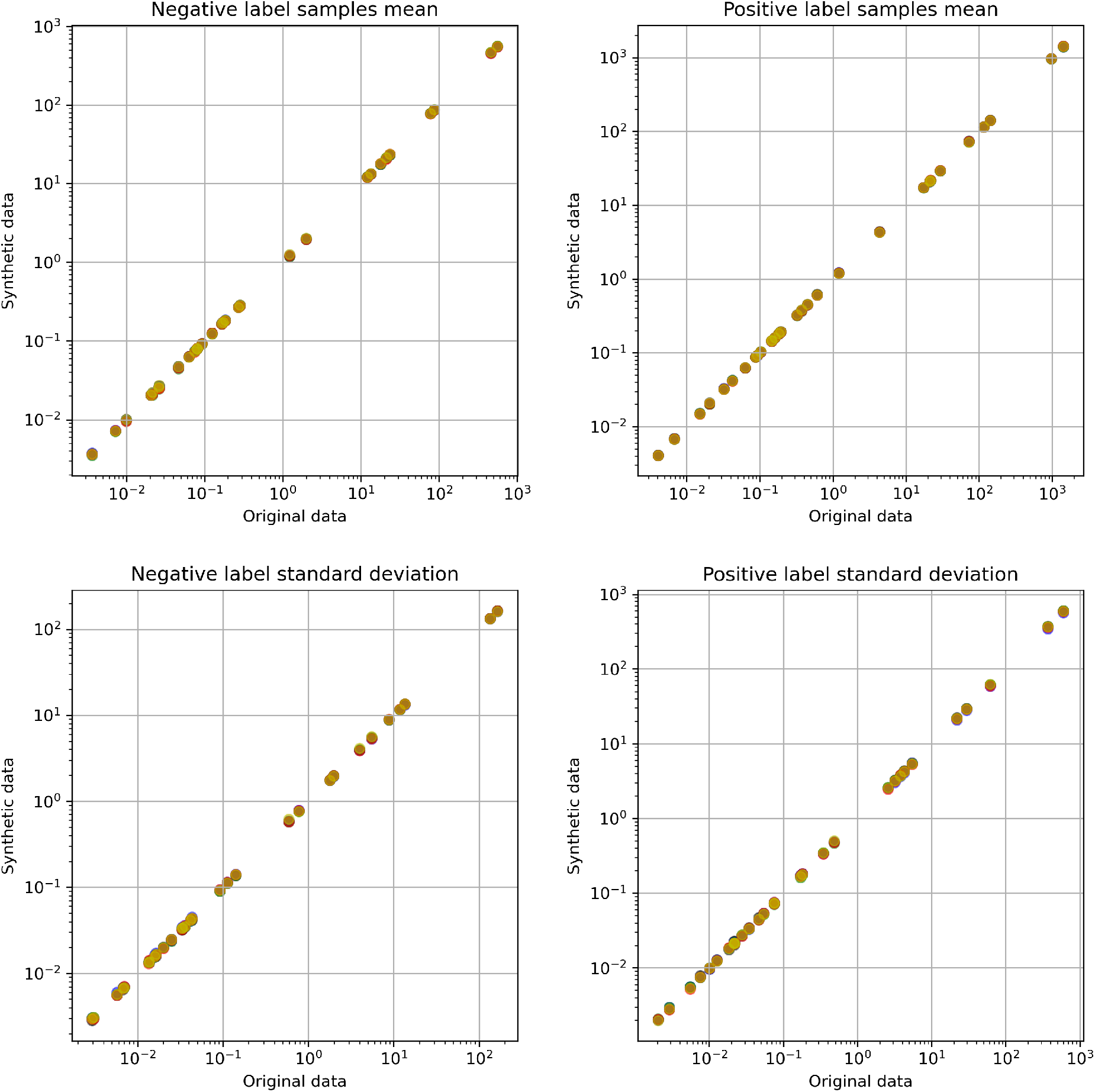
Comparison of mean and standard deviations of features between original and synthetic Wisconsin Breast Cancer data, with synthetic data produced using a Principal Component based approach. Different colours represent five alaternative model runs.

**Figure 2:**
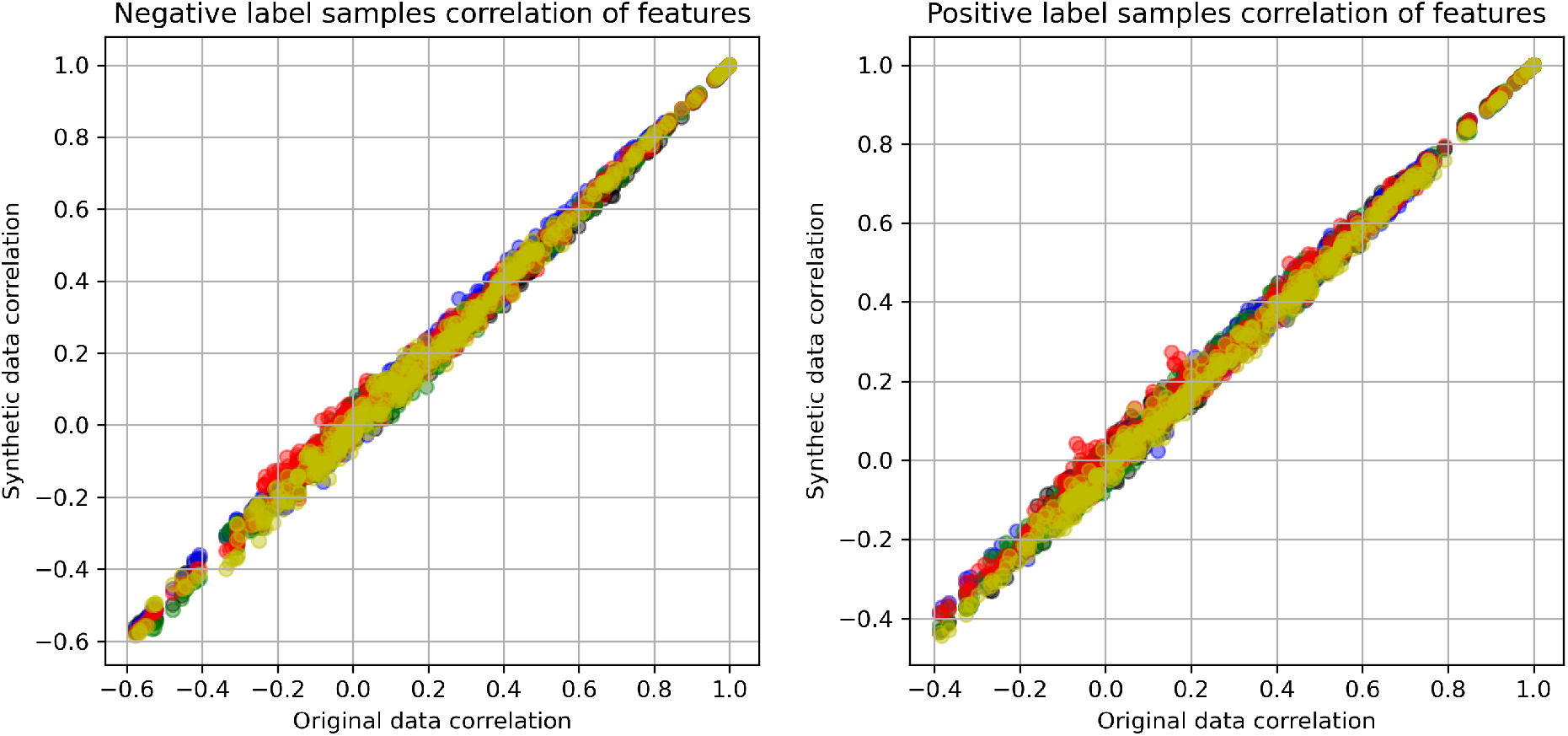
Comparison of correlation between all features in original and synthetic Wisconsin Breast Cancer data with synthetic data produced using a Principal Component based approach. Different colours represent five alaternative model runs.

**Figure 3:**
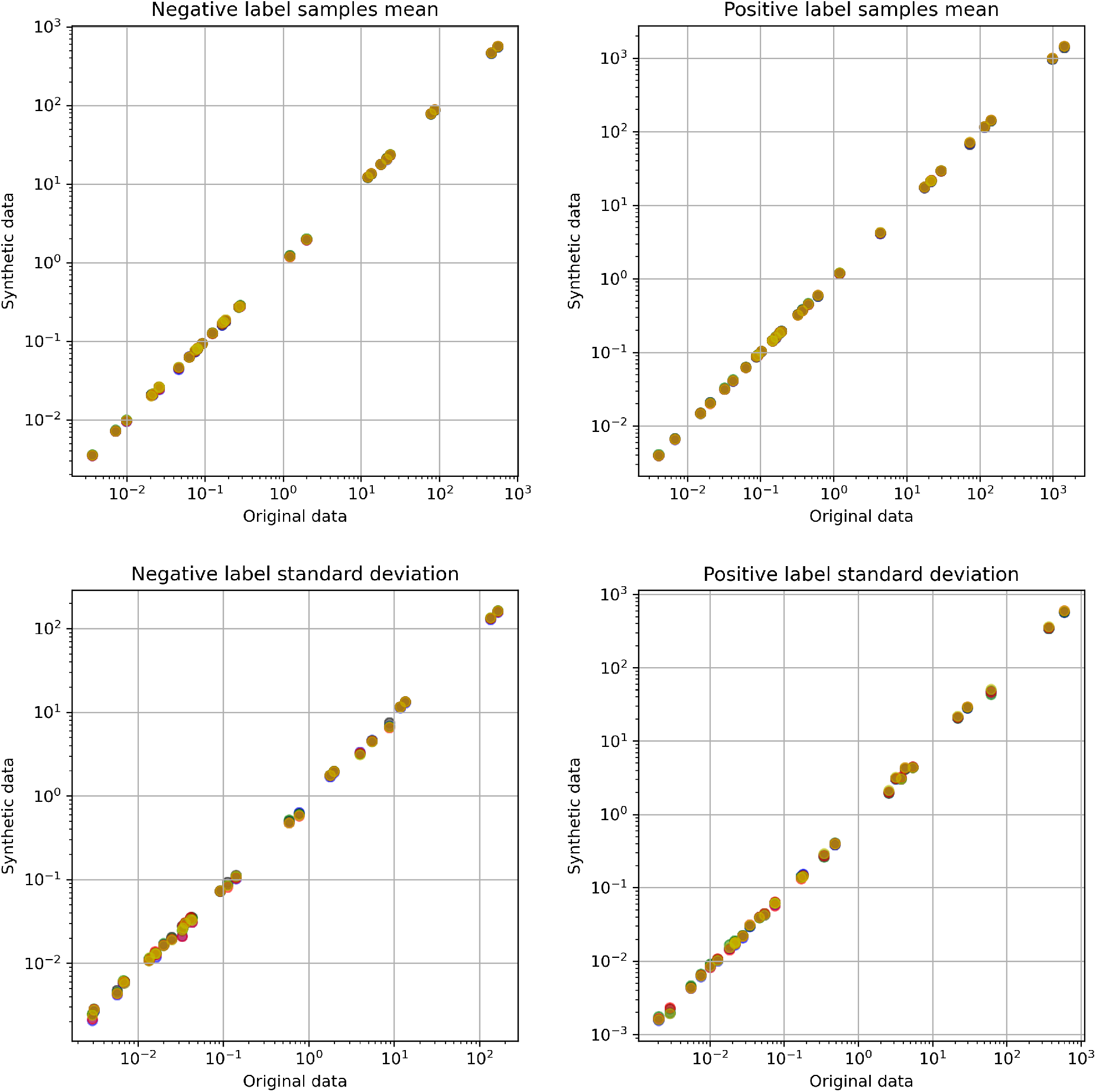
Comparison of mean and standard deviations of features between original and synthetic Wisconsin Breast Cancer data, with synthetic data produced using SMOTE. Different colours represent five alaternative model runs.

**Figure 4:**
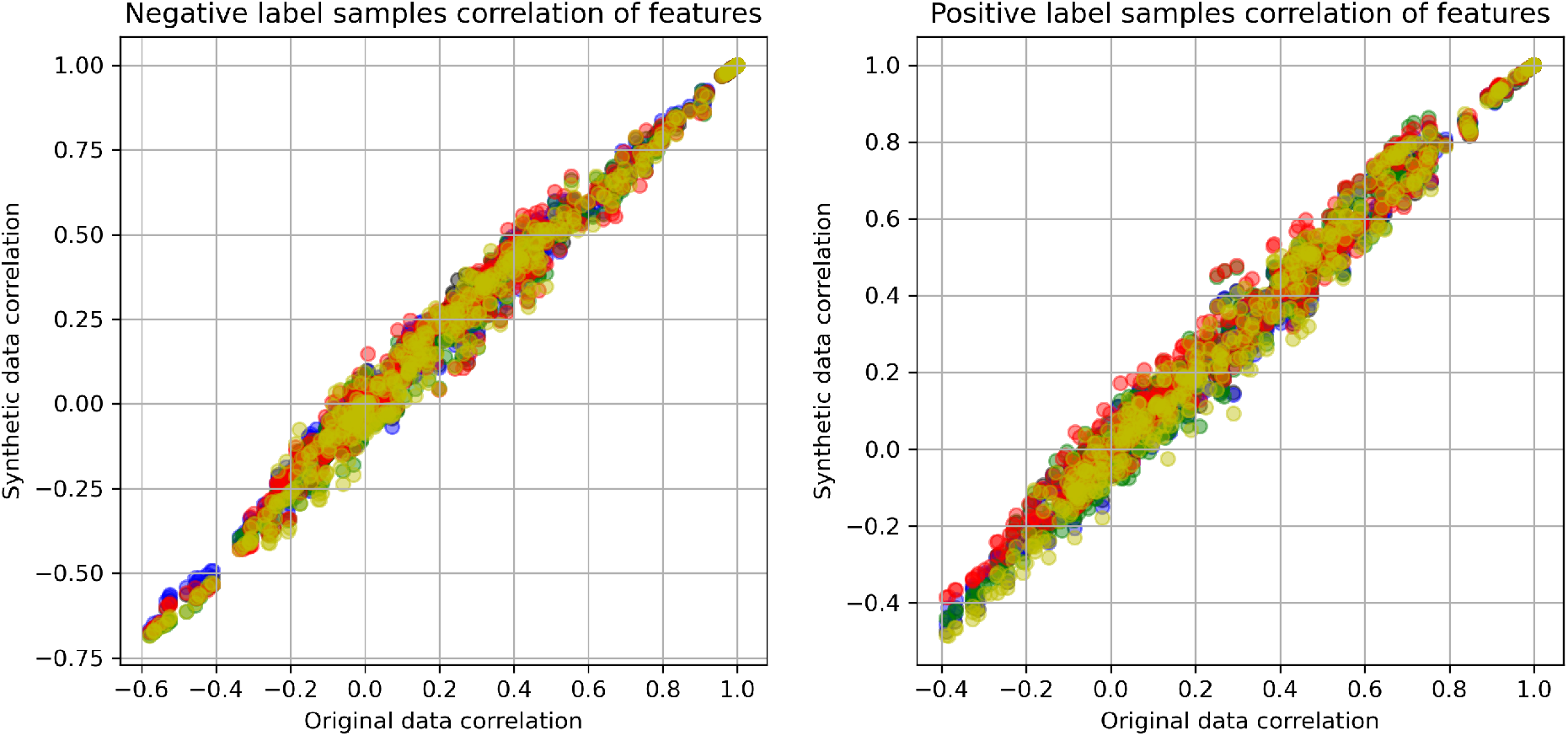
Comparison of correlation between all features in original and synthetic Wisconsin Breast Cancer data with synthetic data produced using SMOTE. Different colours represent five alaternative model runs.

**Figure 5:**
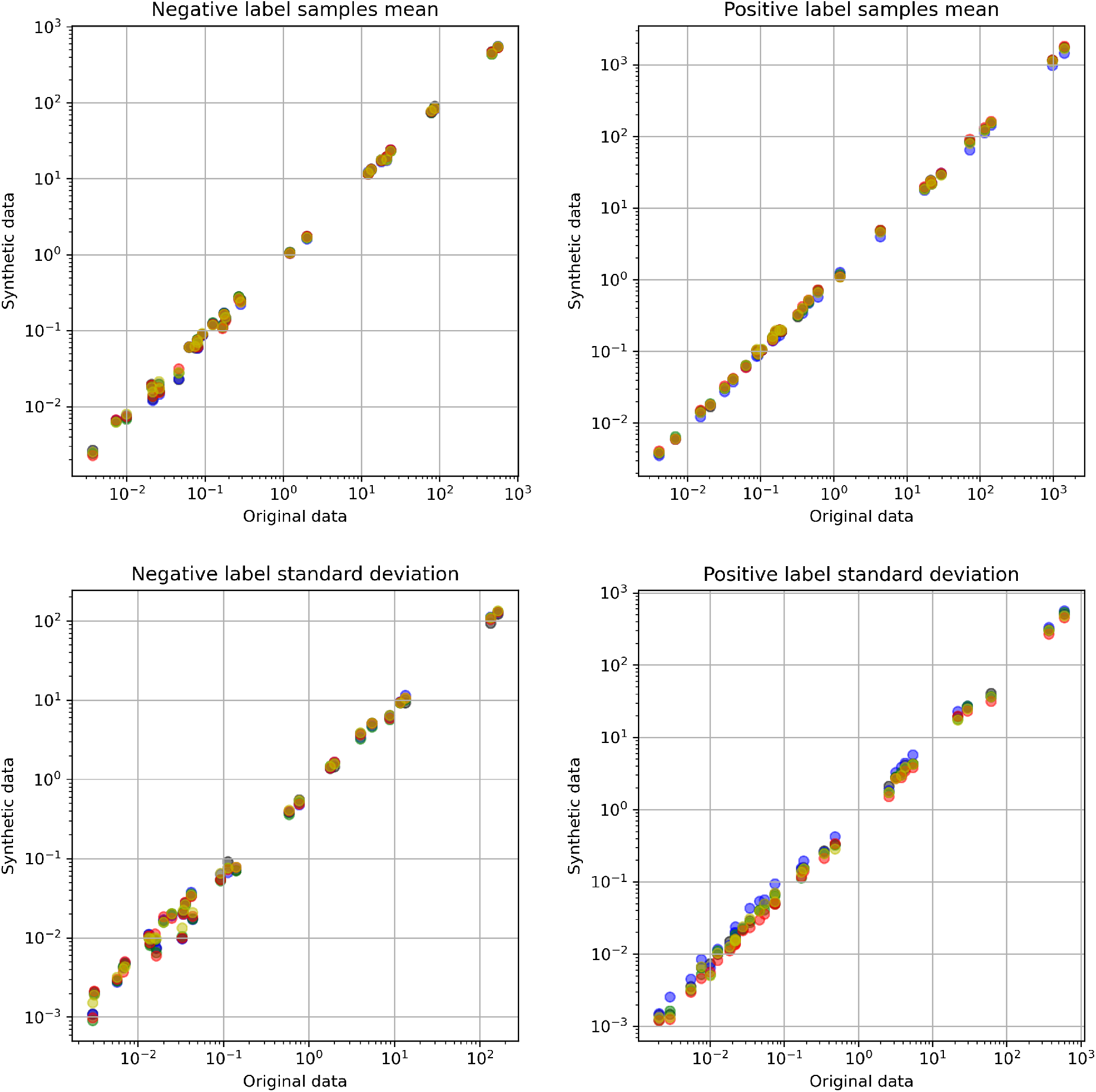
Comparison of mean and standard deviations of features between original and synthetic Wisconsin Breast Cancer data, with synthetic data produced using a Generate Adversarial Network. Different colours represent five alaternative model runs.

**Figure 6:**
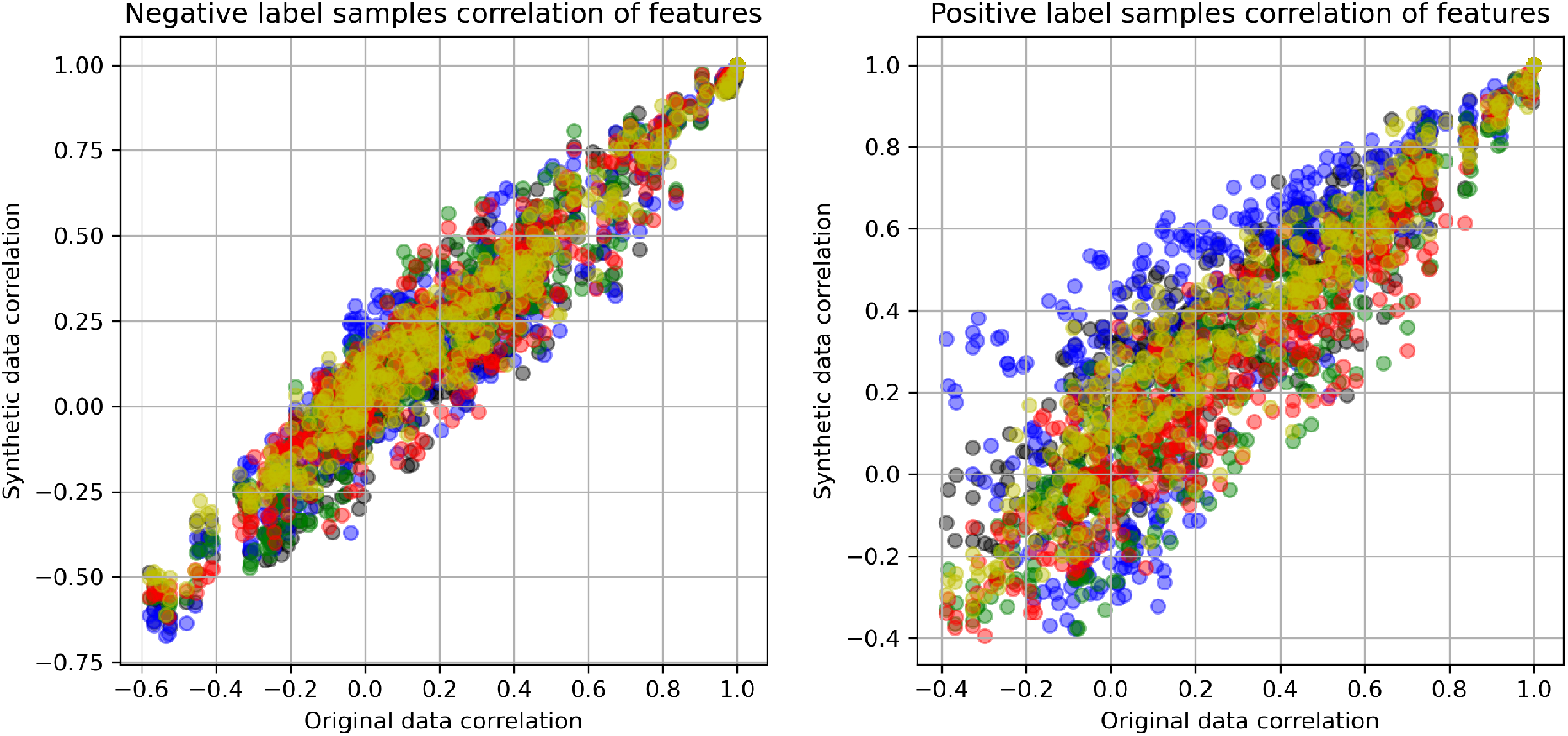
Comparison of correlation between all features in original and synthetic Wisconsin Breast Cancer data with synthetic data produced using a Generate Adversarial Network. Different colours represent five alaternative model runs.

**Figure 7:**
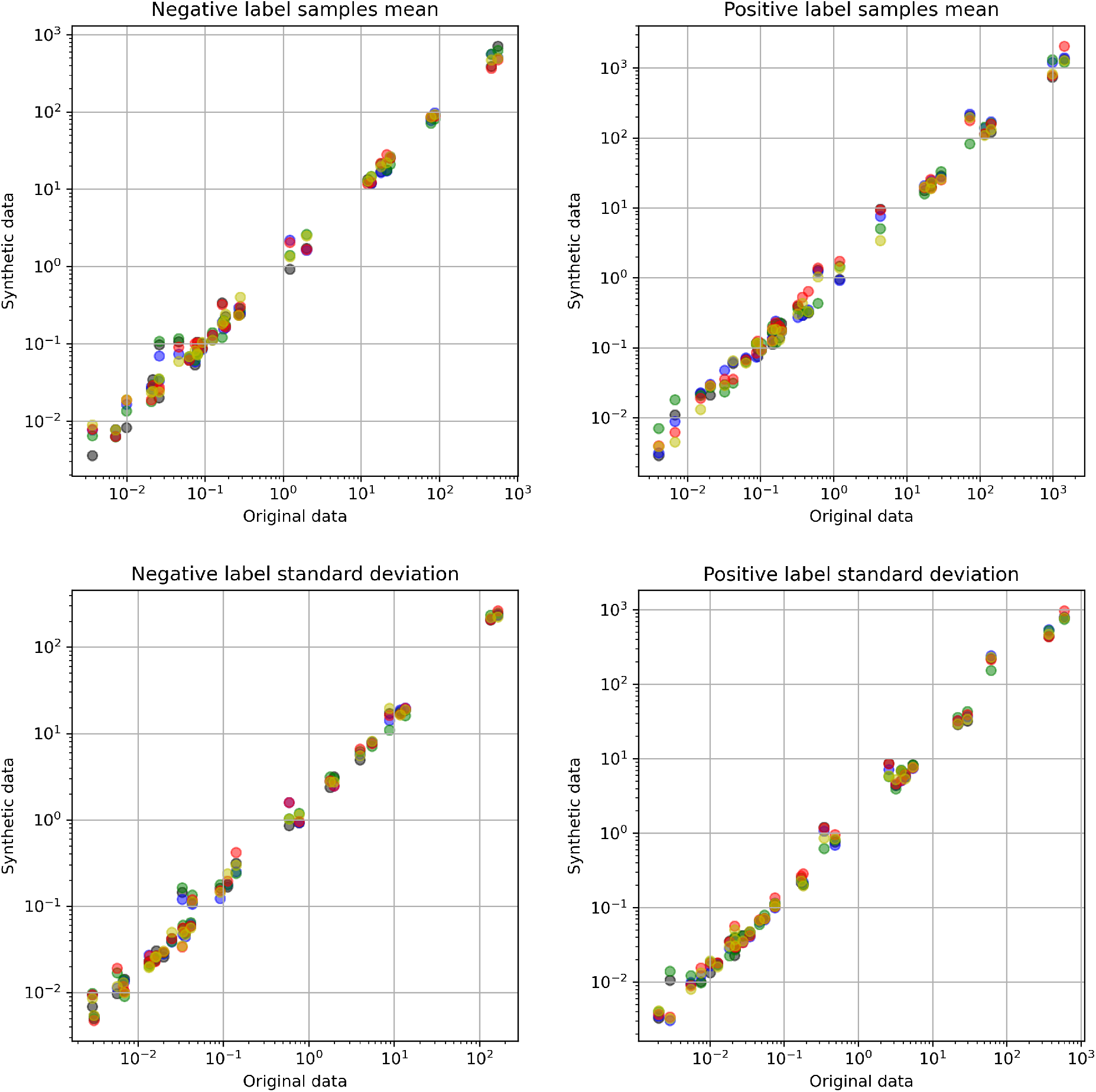
Comparison of mean and standard deviations of features between original and synthetic Wisconsin Breast Cancer data, with synthetic data produced using CT-GAN. Different colours represent five alaternative model runs.

**Figure 8:**
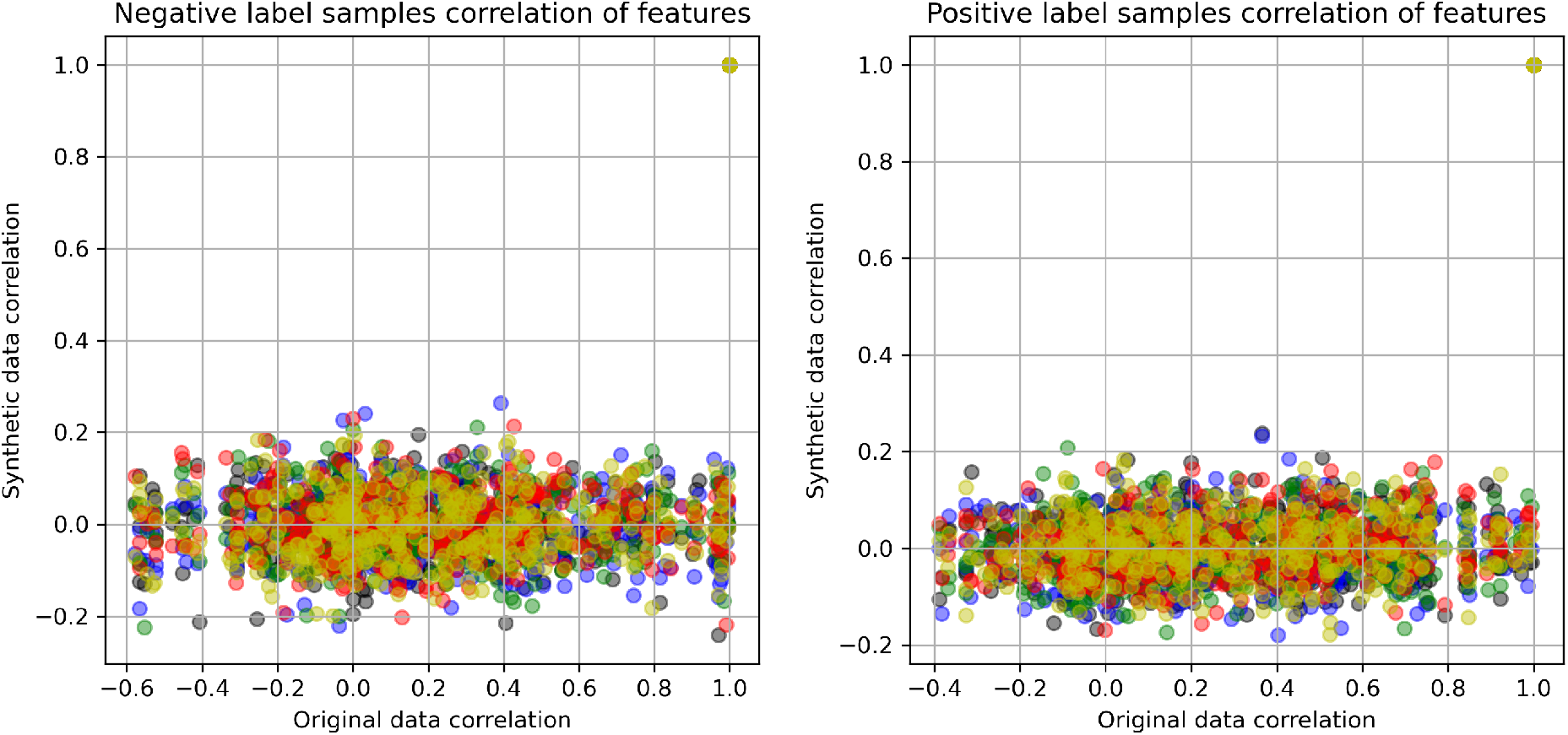
Comparison of correlation between all features in original and synthetic Wisconsin Breast Cancer data with synthetic data produced using CT-GAN. Different colours represent five alaternative model runs.

**Figure 9:**
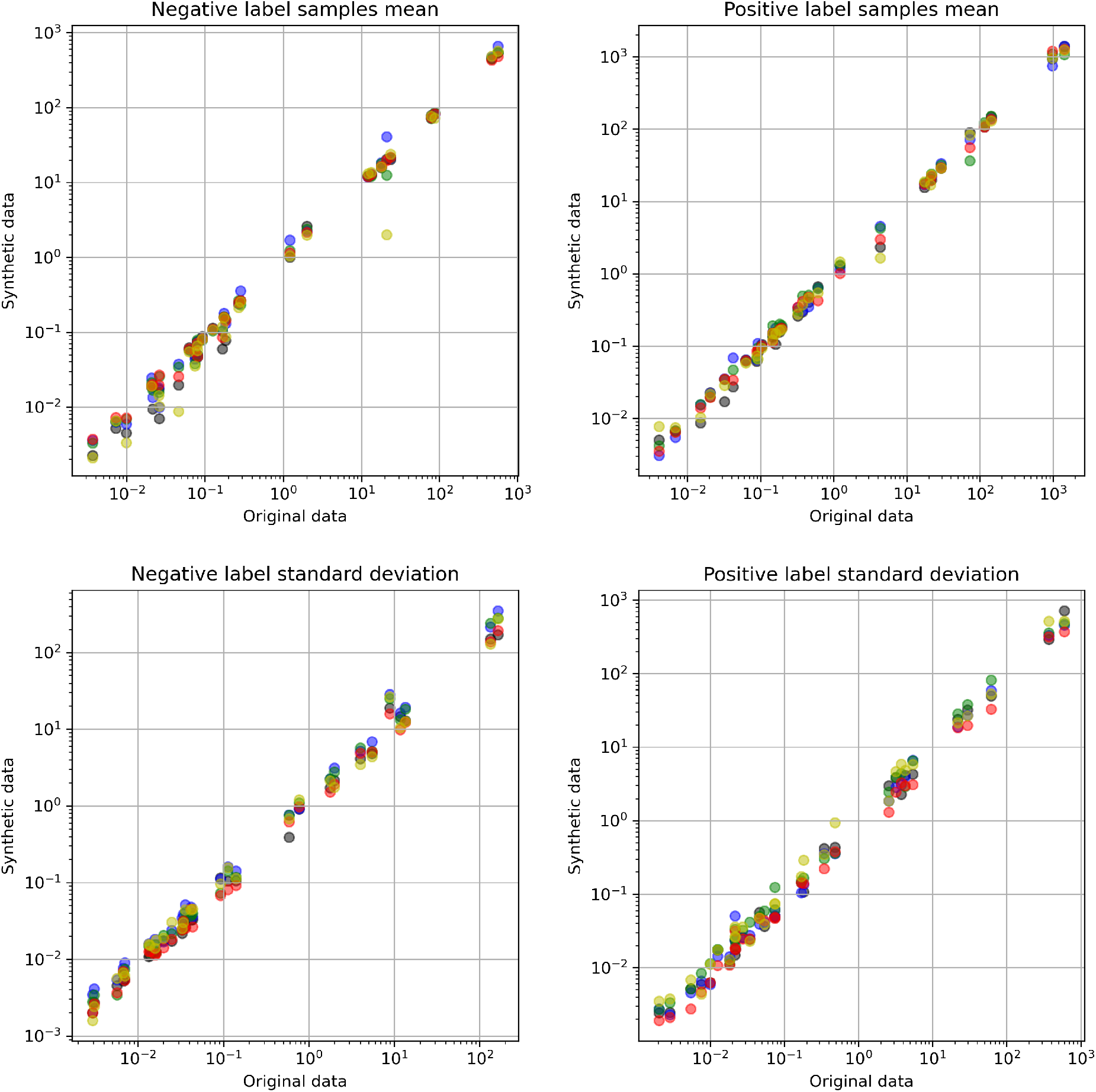
Comparison of mean and standard deviations of features between original and synthetic Wisconsin Breast Cancer data, with synthetic data produced using a Variational Auto Encoder. Different colours represent five alaternative model runs.

**Figure 10:**
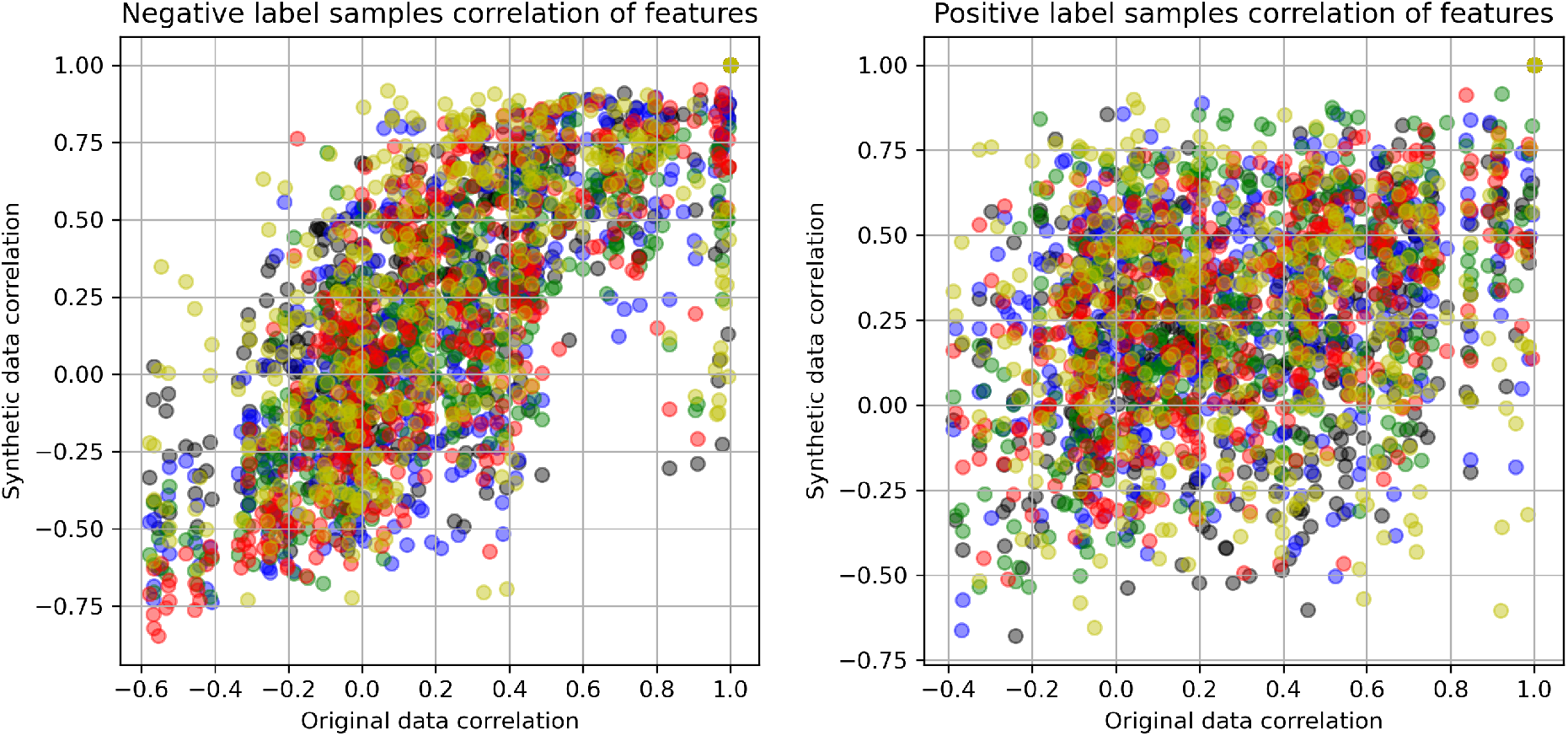
Comparison of correlation between all features in original and synthetic Wisconsin Breast Cancer data with synthetic data produced using a Variational Auto Encoder. Different colours represent five alaternative model runs.

### B Stroke thrombolysis pathway data set

Figures 11 to 20 show a) a comparison of means and standard deviations between the original and synthetic data sets, and b) correlation between all features in original and synthetic data sets. The figures show five synthetic runs for each method.

**Figure 11:**
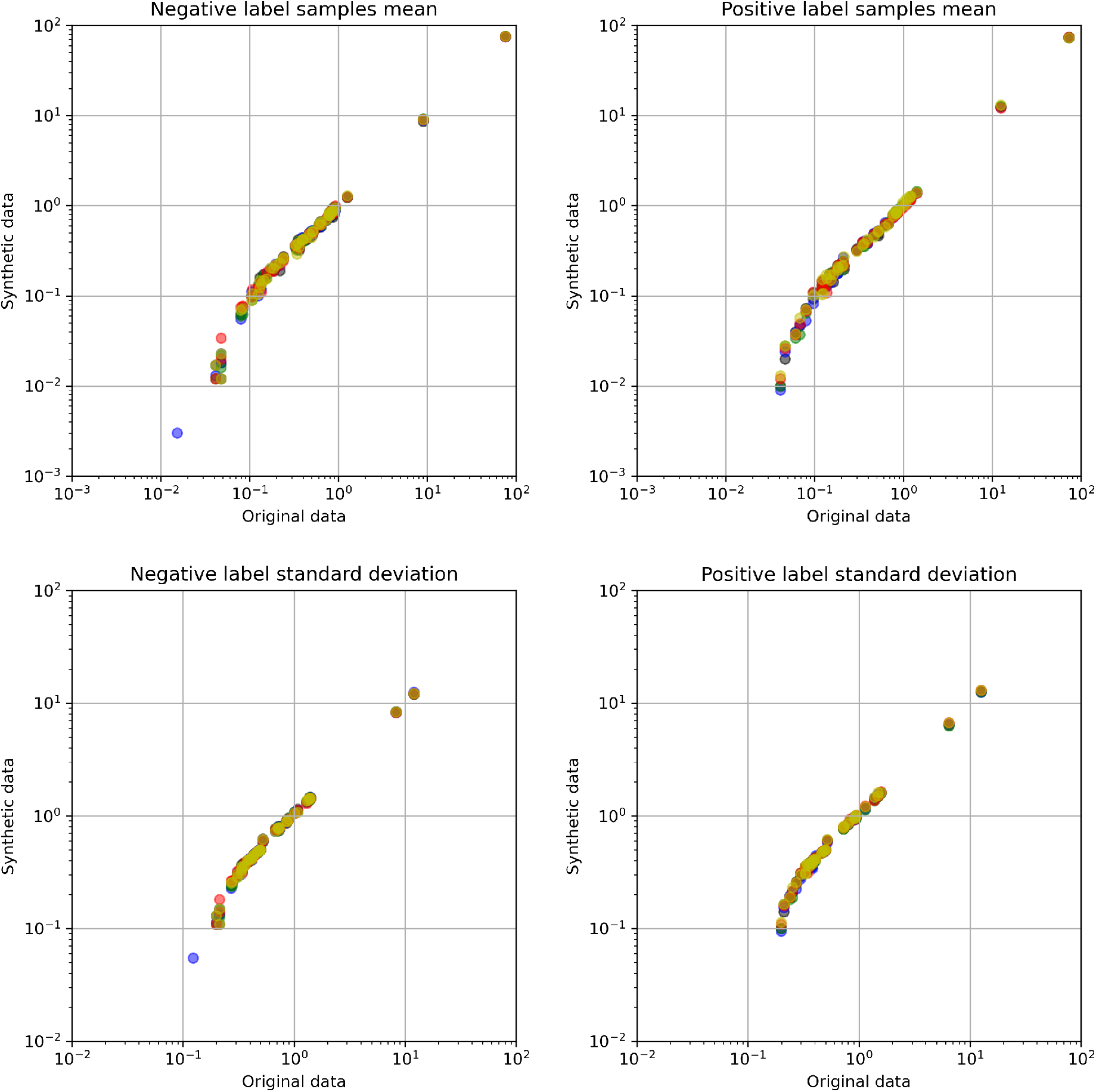
Comparison of mean and standard deviations of features between original and synthetic stroke thrombolysis pathway data, with synthetic data produced using a Principal Component based approach. Different colours represent five alaternative model runs.

**Figure 12:**
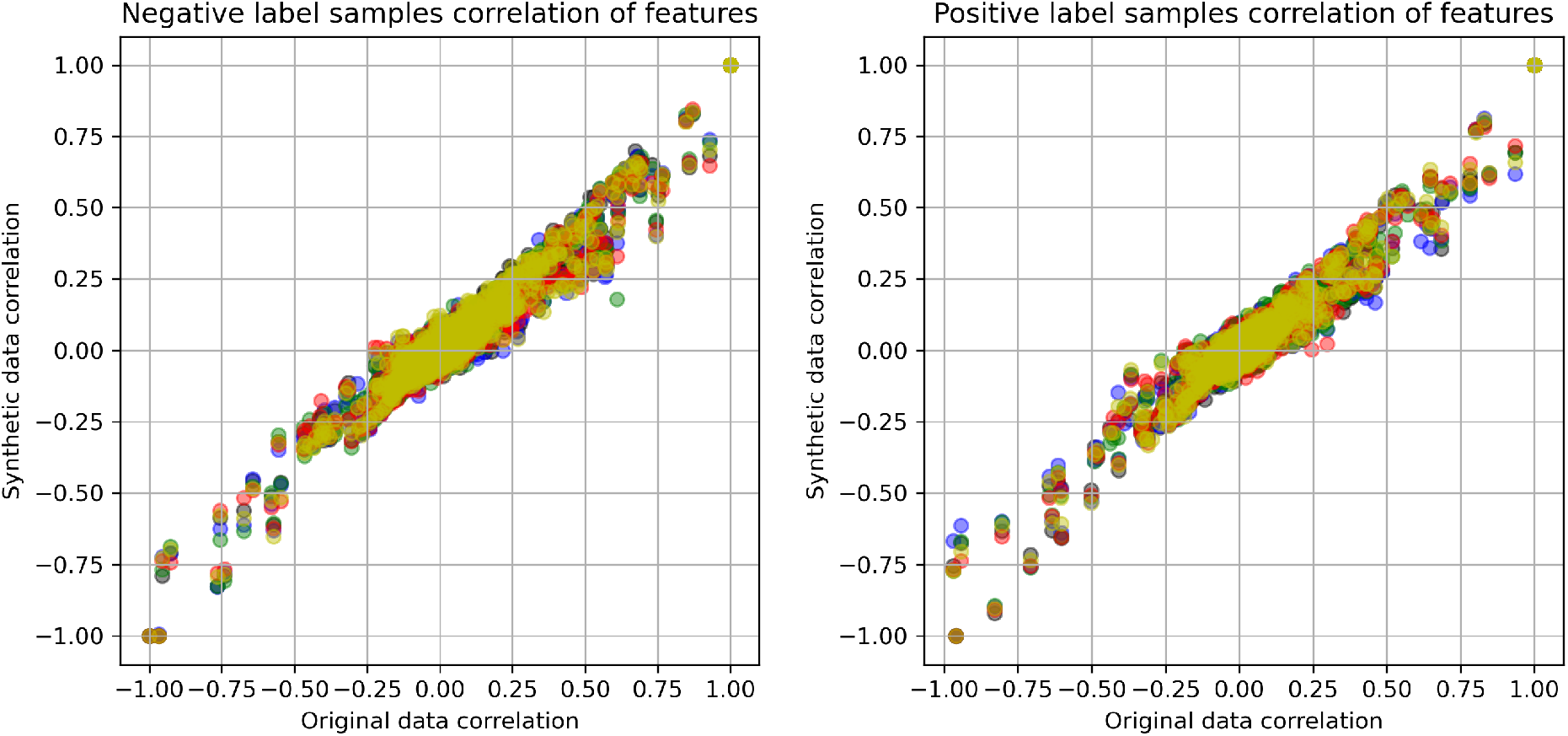
Comparison of correlation between all features in original and synthetic stroke thrombolysis pathway data with synthetic data produced using a Principal Component based approach. Different colours represent five alaternative model runs.

**Figure 13:**
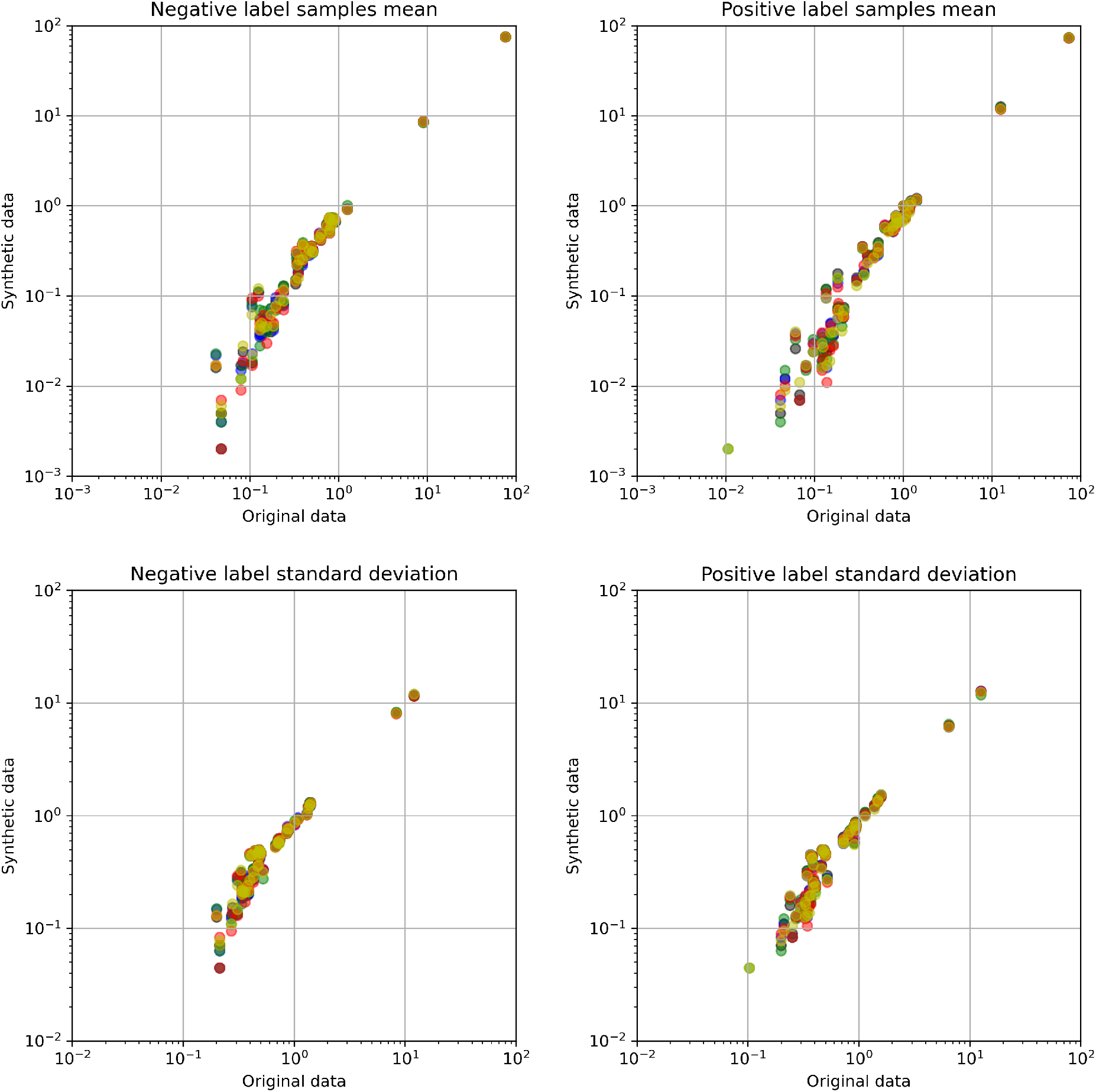
Comparison of mean and standard deviations of features between original and synthetic stroke thrombolysis pathway data, with synthetic data produced using SMOTE. Different colours represent five alaternative model runs.

**Figure 14:**
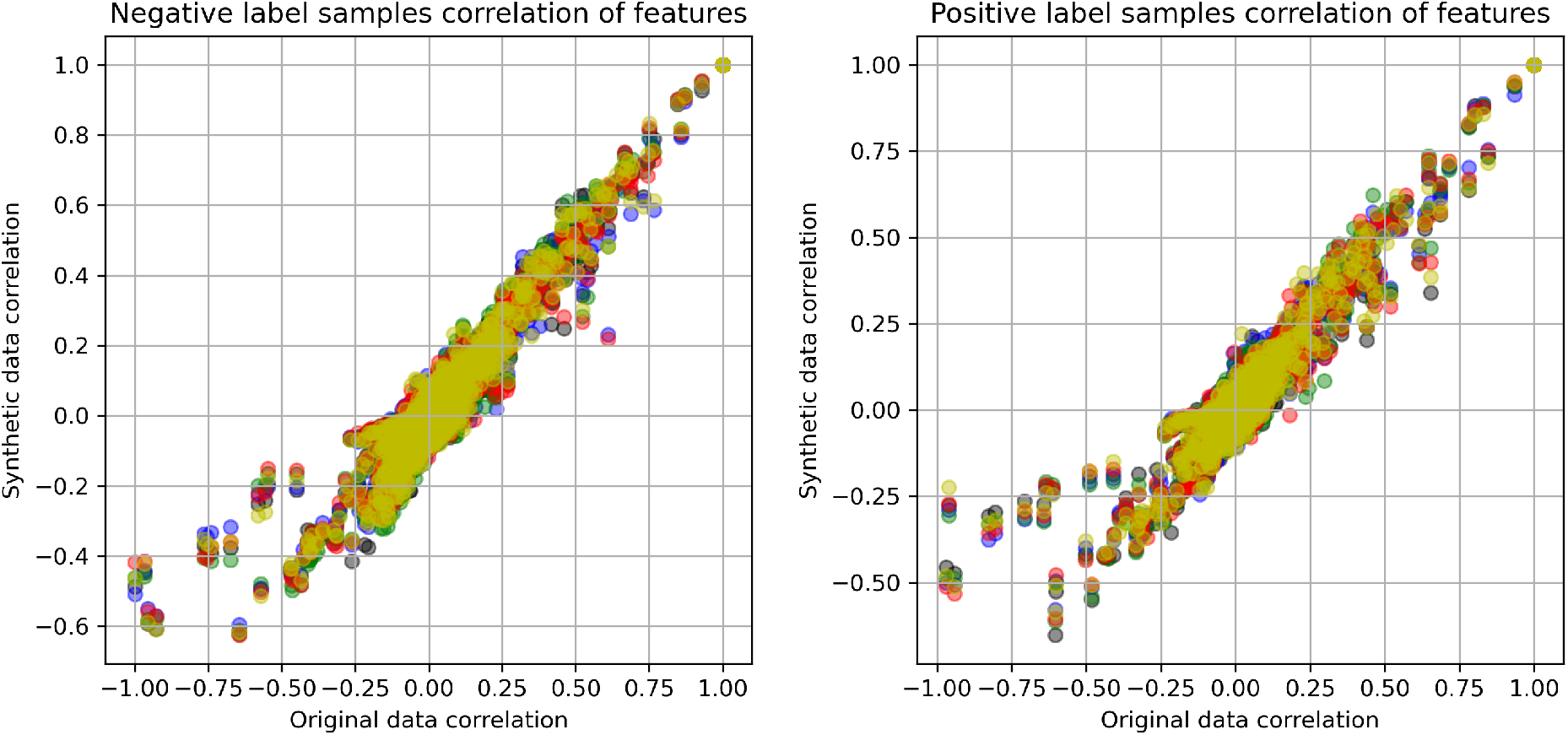
Comparison of correlation between all features in original and synthetic stroke thrombolysis pathway data with synthetic data produced using SMOTE. Different colours represent five alaternative model runs.

**Figure 15:**
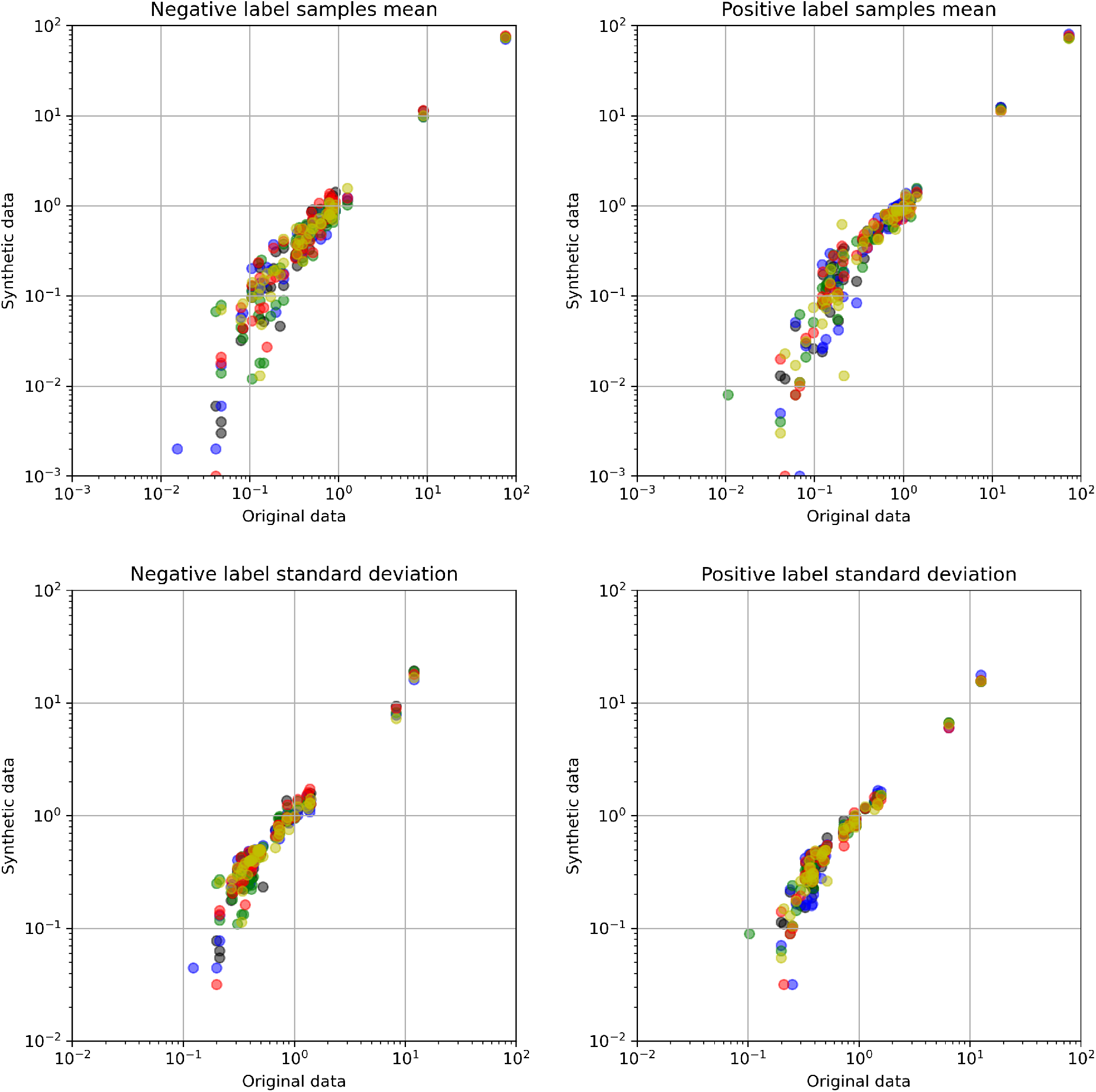
Comparison of mean and standard deviations of features between original and synthetic stroke thrombolysis pathway data, with synthetic data produced using a Generate Adversarial Network. Different colours represent five alaternative model runs.

**Figure 16:**
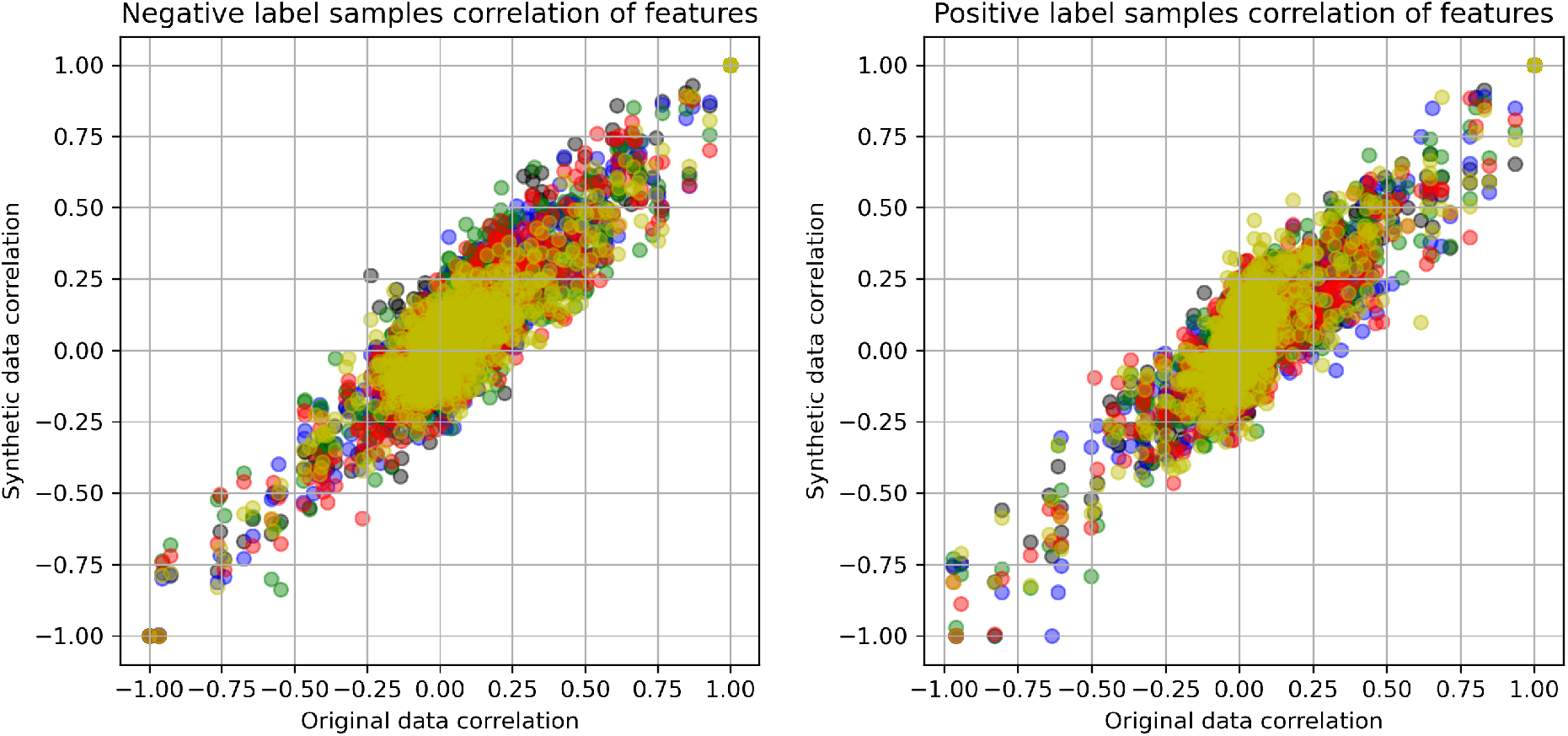
Comparison of correlation between all features in original and synthetic stroke thrombolysis pathway data with synthetic data produced using a Generate Adversarial Network. Different colours represent five alaternative model runs.

**Figure 17:**
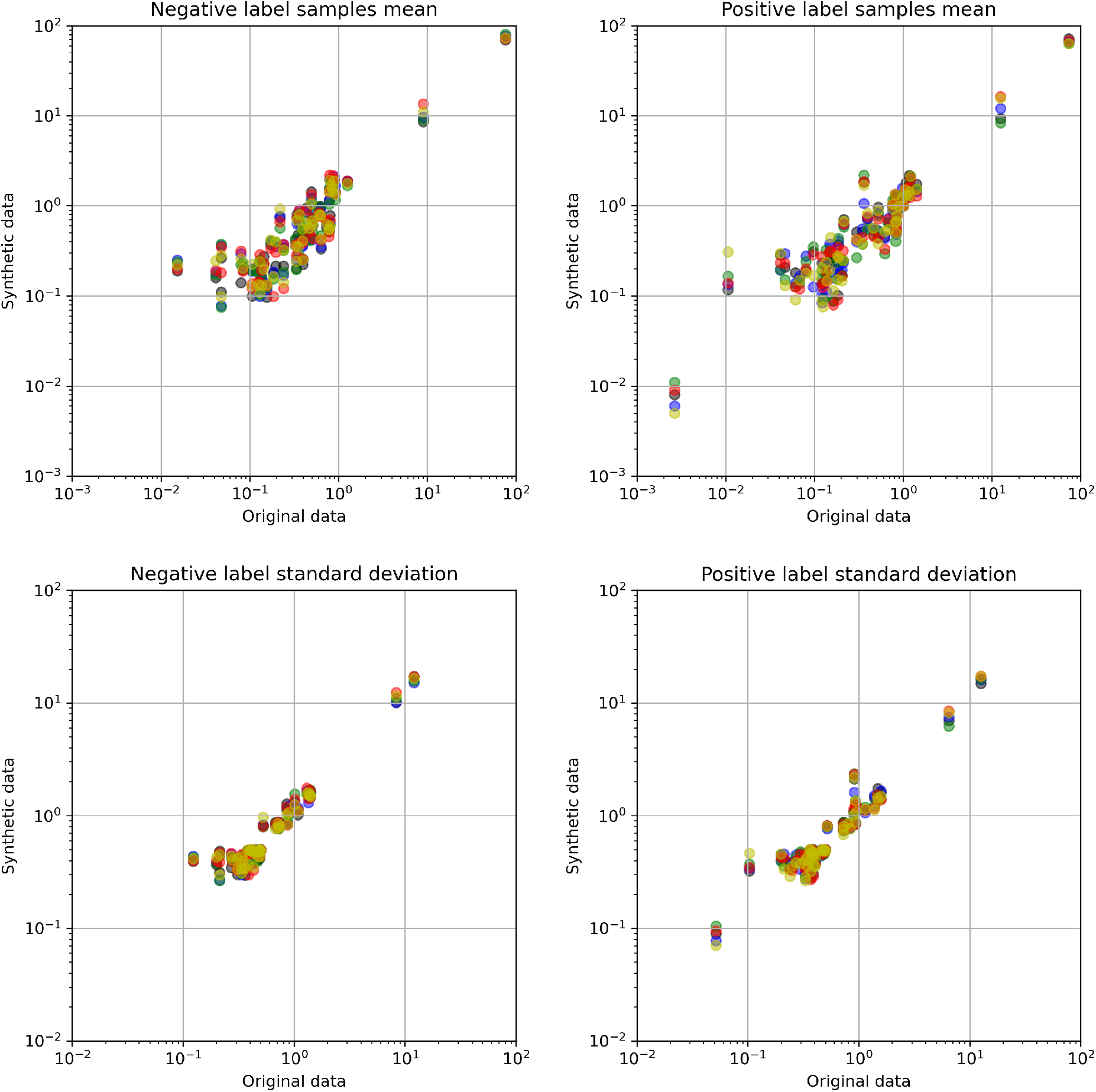
Comparison of mean and standard deviations of features between original and synthetic stroke thrombolysis pathway data, with synthetic data produced using CT-GAN. Different colours represent five alaternative model runs.

**Figure 18:**
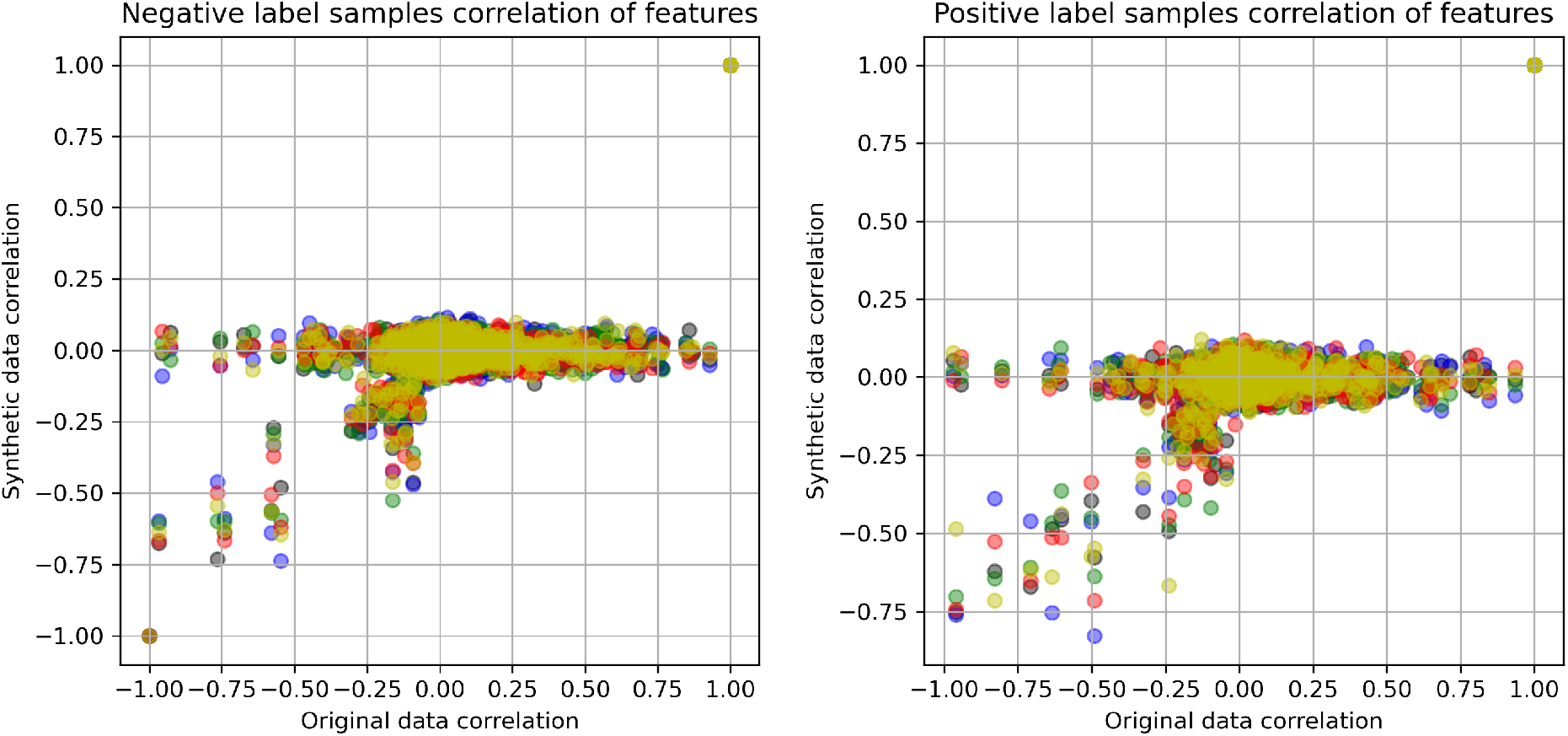
Comparison of correlation between all features in original and synthetic stroke thrombolysis pathway data with synthetic data produced using CT-GAN. Different colours represent five alaternative model runs.

**Figure 19:**
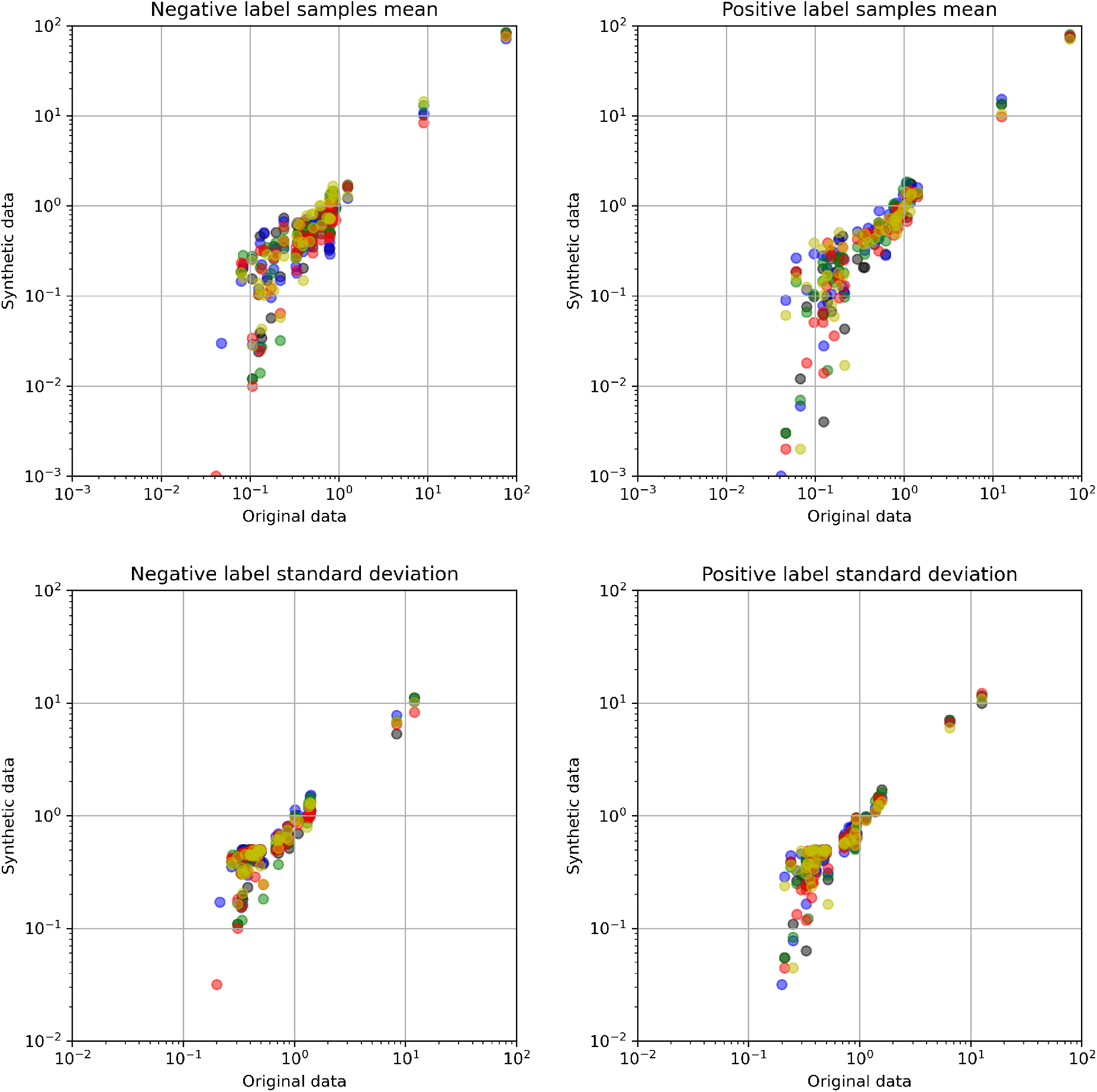
Comparison of mean and standard deviations of features between original and synthetic stroke thrombolysis pathway data, with synthetic data produced using a Variational Auto Encoder. Different colours represent five alaternative model runs.

**Figure 20:**
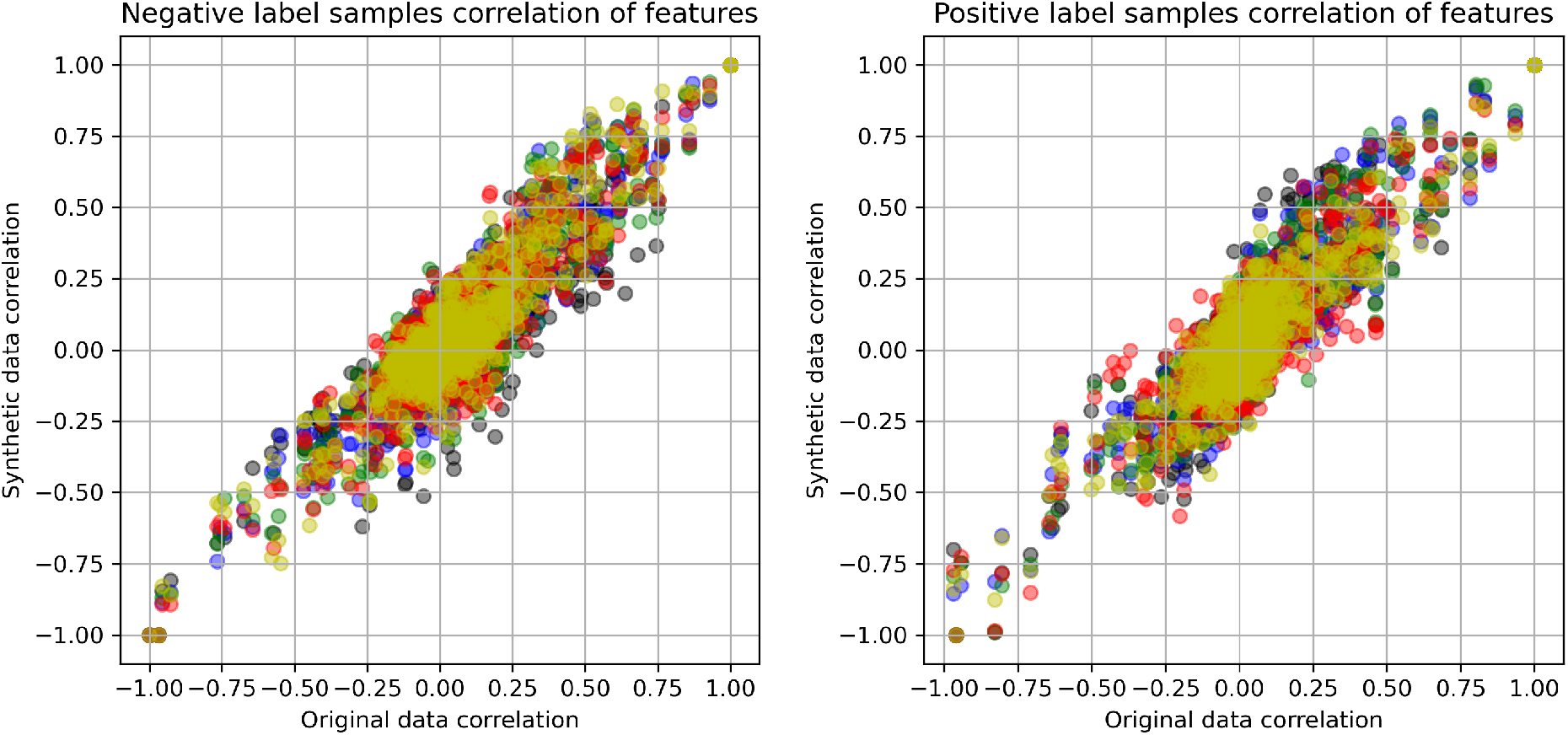
Comparison of correlation between all features in original and synthetic stroke thrombolysis pathway data with synthetic data produced using a Variational Auto Encoder. Different colours represent five alaternative model runs.

## Notes

### Competing Interest Statement

The authors have declared no competing interest.

### Funding Statement

This article presents independent research funded by the National Institute for Health Research (NIHR) Applied Research Collaboration South West Peninsula.

### Author Declarations

No identifiable patient data accessed (public domain anonymous data used). No need to seek approval or exemption.

## References

[1] M. Woelfle, P. Olliaro, and M. H. Todd, “Open science is a research accelerator,” Nature Chemistry, vol. 3, pp. 745–748, Oct. 2011. Number: 10 Publisher: Nature Publishing Group.

[2] C. Dwork, F. McSherry, K. Nissim, and A. Smith, “Calibrating Noise to Sensitivity in Private Data Analysis,” in Theory of Cryptography (S. Halevi and T. Rabin, eds.), Lecture Notes in Computer Science, (Berlin, Heidelberg), pp. 265–284, Springer, 2006.

[3] ZK. P. F.R.S, “On lines and planes of closest fit to systems of points in space,” The London, Edinburgh, and Dublin Philosophical Magazine and Journal of Science, vol. 2, pp. 559–572, Nov. 1901. Publisher: Taylor & Francis eprint: https://doi.org/10.1080/14786440109462720.

[4] N. V. Chawla, K. W. Bowyer, L. O. Hall, and W. P. Kegelmeyer, “SMOTE: Synthetic Minority Over-sampling Technique,” Journal of Artificial Intelligence Research, vol. 16, pp. 321–357, June 2002. 1106.1813.

[5] I. Goodfellow, J. Pouget-Abadie, M. Mirza, B. Xu, D. Warde-Farley, S. Ozair, A. Courville, and Y. Bengio, “Generative Adversarial Nets,” in Advances in Neural Information Processing Systems 27 (Z. Ghahramani, M. Welling, C. Cortes, N. D. Lawrence, and K. Q. Weinberger, eds.), pp. 2672–2680, Curran Associates, Inc., 2014.

[6] L. Xu, M. Skoularidou, A. Cuesta-Infante, and K. Veeramachaneni, “Modeling Tabular data using Conditional GAN,” arXiv:1907.00503 [cs, stat], Oct. 2019. 1907.00503.

[7] D. P. Kingma and M. Welling, “Auto-Encoding Variational Bayes,” arXiv:1312.6114 [cs, stat], Dec. 2013. 1312.6114.

[8] M. A. Kramer, “Nonlinear principal component analysis using autoassociative neural networks,” AIChE Journal, vol. 37, no. 2, pp. 233–243, 1991. eprint: https://aiche.onlinelibrary.wiley.com/doi/pdf/10.1002/aic.690370209.

[9] N. Street, W. Wolberg, and O. Mangasarian, “Nuclear Feature Extraction For Breast Tumor Diagnosis,” Proc. Soc. Photo-Opt. Inst. Eng., vol. 1993, Jan. 1999.

[10] M. Allen, K. Pearn, T. Monks, B. D. Bray, R. Everson, A. Salmon, M. James, and K. Stein, “Can clinical audits be enhanced by pathway simulation and machine learning? An example from the acute stroke pathway,” BMJ Open, vol. 9, p. e028296, Sept. 2019. Publisher: British Medical Journal Publishing Group Section: Health services research.

[11] G. Van Rossum and F. L. Drake Jr, Python reference manual. Centrum voor Wiskunde en Informatica Amsterdam, 1995.

[12] F. Pedregosa, G. Varoquaux, A. Gramfort, V. Michel, B. Thirion, O. Grisel, M. Blondel, P. Prettenhofer, R. Weiss, V. Dubourg, J. Vanderplas, A. Passos, D. Cournapeau, M. Brucher, M. Perrot, and E. Duchesnay, “Scikit-learn: Machine Learning in Python,” Journal of Machine Learning Research, vol. 12, no. 85, pp. 2825–2830, 2011.

[13] G. Lemaitre, F. Nogueira, and C. K. Aridas, “Imbalanced-learn: A Python Toolbox to Tackle the Curse of Imbalanced Datasets in Machine Learning,” arXiv:1609.06570 [cs], Sept. 2016. arXiv: 1609.06570.

[14] A. Paszke, S. Gross, F. Massa, A. Lerer, J. Bradbury, G. Chanan, T. Killeen, Z. Lin, N. Gimelshein, L. Antiga, A. Desmaison, A. Kopf, E. Yang, Z. DeVito, M. Raison, A. Tejani, S. Chilamkurthy, B. Steiner, L. Fang, J. Bai, and S. Chintala, “PyTorch: An Imperative Style, High-Performance Deep Learning Library,” in Advances in Neural Information Processing Systems 32 (H. Wallach, H. Larochelle, A. Beygelzimer, F. d. Alché-Buc, E. Fox, and R. Garnett, eds.), pp. 8024–8035, Curran Associates, Inc., 2019.

[15] M. Abadi, A. Agarwal, P. Barham, E. Brevdo, Z. Chen, C. Citro, G. S. Corrado, A. Davis, J. Dean, M. Devin, S. Ghemawat, I. Goodfellow, A. Harp, G. Irving, M. Isard, Y. Jia, R. Jozefowicz, L. Kaiser, M. Kudlur, J. Levenberg, D. Mané, R. Monga, S. Moore, D. Murray, C. Olah, M. Schuster, J. Shlens, B. Steiner, I. Sutskever, K. Talwar, P. Tucker, V. Vanhoucke, V. Vasudevan, F. Viégas, O. Vinyals, P. Warden, M. Wattenberg, M. Wicke, Y. Yu, and X. Zheng, “TensorFlow: Large-scale machine learning on heterogeneous systems,” 2015. Software available from tensorflow.org.

[16] C. R. Harris, K. J. Millman, S. J. van der Walt, R. Gommers, P. Virtanen, D. Cournapeau, E. Wieser, J. Taylor, S. Berg, N. J. Smith, R. Kern, M. Picus, S. Hoyer, M. H. van Kerkwijk, M. Brett, A. Haldane, J. F. del Río, M. Wiebe, P. Peterson, P. Gérard-Marchant, K. Sheppard, T. Reddy, W. Weckesser, H. Abbasi, C. Gohlke, and T. E. Oliphant, “Array programming with NumPy,” Nature, vol. 585, pp. 357–362, Sept. 2020. Number: 7825 Publisher: Nature Publishing Group.

[17] Wes McKinney, “Data Structures for Statistical Computing in Python,” in Proceedings of the 9th Python in Science Conference (Stéfan van der Walt and Jarrod Millman, eds.), pp. 56–61, 2010.

[18] J. D. Hunter, “Matplotlib: A 2d graphics environment,” Computing in Science & Engineering, vol. 9, no. 3, pp. 90–95, 2007.

[19] M. Arjovsky, S. Chintala, and L. Bottou, “Wasserstein GAN,” arXiv:1701.07875 [cs, stat], Dec. 2017. arXiv: 1701.07875.

